# High seroreactivity against SARS-CoV-2 Spike epitopes in a pre SARS-CoV-2 cohort: implications for antibody testing and vaccine design

**DOI:** 10.1101/2020.05.18.20105189

**Authors:** Kaia Palm, Mariliis Jaago, Annika Rähni, Nadežda Pupina, Arno Pihlak, Helle Sadam, Annela Avarlaid, Anu Planken, Margus Planken, Liina Haring, Eero Vasar, Miljana Baćević, France Lambert, Eija Kalso, Pirkko Pussinen, Pentti J. Tienari, Antti Vaheri, Dan Lindholm, Tõnis Timmusk, Amir M Ghaemmaghami

**Affiliations:** Protobios Llc, Tallinn, Estonia. Electronic address; Department of Chemistry and Biotechnology, Tallinn University of Technology, Tallinn, Estonia; North Estonia Medical Centre Foundation, Tallinn, Estonia; Institute of Clinical Medicine, University of Tartu, Psychiatry Clinic of Tartu University Hospital, Estonia; Department of Physiology, Institute of Biomedicine and Translational Medicine, University of Tartu, Tartu, Estonia; Center of Excellence for Genomics and Translational Medicine, University of Tartu, Tartu, Estonia; Dental Biomaterial Research Unit (d-BRU), Faculty of Medicine, University of Liege, Liege, Belgium; Department of Periodontology and Oral Surgery, Faculty of Medicine, University of Liege, Belgium; Department of Anaesthesiology, Intensive Care and Pain Medicine, Helsinki Uiversity Hospital and Department of Pharmacology and SleepWell Research Programme, University of Helsinki, Helsinki, Finland; Oral and Maxillofacial Diseases, University of Helsinki and Helsinki University Hospital, Helsinki, Finland; Department of Neurology, Neurocenter, Helsinki University Hospital, and Translational Immunology Research Program, University of Helsinki, Helsinki, Finland; Department of Virology, Medicum, University of Helsinki, Finland; Department of Biochemistry and Developmental Biology, Faculty of Medicine, University of Helsinki, Helsinki, Finland; Minerva Foundation Institute for Medical Research, Helsinki, Finland; Immunology and Immuno-Bioengineering Group, School of Life Science, Faculty of Medicine and Health Sciences, University of Nottingham, Nottingham, United Kingdom. Electronic address

**Keywords:** SARS-CoV-2, immunoprfiling, antibody, seroreactivity, COVID-19, antigenic sin

## Abstract

Little is known about the quality of polyclonal antibody responses in COVID-19 patients, and how it correlates with disease severity or patients’ prior exposure to other pathogens. The whole polyclonal antibody repertoire in a retrospective cohort of 538 individuals was mapped against SARS-CoV-2 spike (S) glycoprotein, the main target of antibody immune responses in SARS-CoV-2 infection. Bioinformatic predictions identified 15 major B cell epitopes for S of SARS-CoV-2. Several epitopes localised in RBD of S including those spanning the ACE2-binding site, the highly conserved cryptic epitope of the neutralizing antibody of SARS-CoV, and fusion/entry domains of HR1 and HR2 of S protein of SARS-CoV-2. Intriguingly, some of these epitopes have cross-reactivity to antigens of common pathogens, potentially affecting SARS-CoV-2 infection outcome. High level of anti-Spike SARS-CoV-2 seroreactivity in populations with no history of exposure to SARS-CoV-2 is of clinical relevance and could underpin better understanding of COVID-19 pathophysiology in different populations and provide a blueprint for design of effective vaccines and developing better strategies for antibody testing.

## Intorduction

COVID-19 pandemic has caused unprecedented health and economic challenges. A large proportion of people infected by SARS-CoV-2 can be asymptomatic^1^, whilst in those with symptoms, the disease can cause a spectrum of clinical manifestations from mild upper respiratory tract illness to cytokine storm and severe pneumonia leading to respiratory failure and death^2^. Older age and a number of co-morbidities (e.g. diabetes, cardiovascular diseases)^2-7^ have been associated with the development of more severe disease however the underlying mechanisms of poor clinical outcome have remained elusive. Different patterns of immune response in COVID-19 patients are linked both to full recovery (e.g. emergence of antibody secreting cells and SARS-CoV-2 binding antibodies) and to poor clinical outcome (immune dysregulation and prolonged inflammation)^8^ and death. However, the factors driving development of beneficiary versus harmful immune responses are unclear. One of the key features of the human immune system is its memory. First exposure to many of the common human viruses usually occurs during childhood or adolescence that shapes our immune memory and the immune response to self and environmental antigens throughout life^9,10^. The immune system’s response to a new pathogen can be influenced by its memory of the response to a variant of that pathogen or a cross-reactive antigen. This is also known as ‘original antigenic sin’ (OAS)^11^, which could lead to either better protection against infection, or to a higher susceptibility and poor clinical outcome. This phenomenon is probably best shown in the context of many viral infections such as influenza and influenza vaccines^12-14^.

In this study, we investigated the presence of seroreactivity to SARS-CoV-2 major antigenic epitopes of spike (S) glycoprotein^15,16^ using a retrospective cohort of pre SARS-CoV2 patients (**Table 1)**. Serum samples from different individuals with varied disease background, with a potential link to COVID-19 severity^2,4-7^, and healthy controls collected between 2011-2019 (see **Methods**) were used to investigate the existence of immunereactivity against SARS-CoV-2 main antigenic epitopes using Mimotope-variation analysis (MVA) (**Methods** and **Supplementary Figure 1**).

**Table 1.** Descriptions of clinical cohorts.

| No | Cohort (abbreviation) | Diagnosis group | Group size (n) | Gender | Age (mean ±SD), years | Ethnicity |
| --- | --- | --- | --- | --- | --- | --- |
| Cardiovascular diseases (with comorbidities including obesity, hypertension, diabetes) |  |  |  |  |  |  |
| 1. | Periodontitis and coronary artery disease (PD-CAD) | No-CAD | 32 | 17M/15F | 60.2 | Finnish |
|  |  | Stable-CAD | 32 | 26M/6F | 64.3 |  |
|  |  | ACS | 32 | 24M/8F | 61.3 |  |
| 2. | Myocardial infarction (MI) | HC | 50 | 20M/30F | 40.8 | Estonian |
|  |  | MI | 50 | 29M/13F/8NA | 66.8 (based on 40 individuals); 10 NA | Estonian |
| 3. | T2D and foot ulcer | T2D | 25 | 21M/4F | 69.0 ± 8.7 | Belgian |
| Diseases with autoimmune component |  |  |  |  |  |  |
| 4. | Multiple sclerosis (MS) | MS | 20 | 4M/16F | 32.3 ± 7.8 | Finnish |
| Cancer |  |  |  |  |  |  |
| 6. | Breast cancer (BC) | Non-NP | 30 | 0M/30F | 57.4 ± 7.8 | Finnish |
|  |  | NP | 27 | 0M/27F | 53.9 ± 6.1 | Finnish |
| Psychiatric diseases |  |  |  |  |  |  |
| 7. | First episode psychosis (FEP) | HC | 30 | 15M/15F | 24.0 ± 6.1 | Estonian |
|  |  | FEP | 30 | 15M/12F/3 NA | 26.0 ± 4.9 (based on 27 individuals); 3 NA | Estonian |
| 8. | Schizophrenia (SZ) | HC | 30 | 12M/18F | 42.1 ± 18.2 | Finnish |
|  |  | SZ | 30 | 12M/18F | 41.9 ± 18.4 | Finnish |

ACS – acute coronary syndrome; NP – neuropathic pain; T2D – type II diabetes with impaired wound healing; HC – healthy control; M – male; F – female; NA – not available

## Results and Discussion

The more abundant and shared peptide antigens of MVA immunoprofiles were clustered into core sequences (motifs) either via group-discriminatory identification (FEP and FEP_HC, SZ and SZ_HC, MS) or non-discriminatory selection (PD-CAD, MI and MI_HC, T2D, BC). Altogether, a set of 22,949 motif sequences were selected, representing the most abundant and common antibody immune response features across age, gender, and different clinical backgrounds (**Supplementary Information**).

Using the data from more prominent immunoprofile features from 538 serum samples, we predicted 15 epitopes within S that were specifically immunodominant (**Figure 1** and **Supplementary Table 1**). Using simulated random alignment data, epitopes predicted by MVA were statistically significant (p<0.05) (**Figure 1**). Of these, 10 overlapped with the 29 CoV-2 reactive B cell epitopes^17^ with average overlap of 60.1% per epitope, due to shorter length of MVA-predicted epitopes. Four of the epitopes (Epitopes 4, 5, 6, and 7, (Ep.4, Ep.5, Ep.6, Ep7)) were mapped to the receptor binding domain (RBD), spanning amino acids 319 to 542 of S that is a crucial antigenic attachment region for binding with ACE2^18^ (the main receptor for host cell infection) (**Figure 1** and **Supplementary Figure 2**). In addition to positioning within the RBD, Ep.5 and Ep.6 overlap with the key amino acids important for binding to human ACE2^18^ (**Figure 1**). Ep.4 and Ep.7 overlap with either the N- or C-terminal region, respectively, of the discontinuous epitope described for CR3022, a neutralizing antibody previously isolated from a convalescent SARS-CoV patient which can also bind the RBD of SARS-CoV-2^18^ (**Figure 1**). Interestingly, Ep.4 includes P384 residue, and Ep.7 contains H519 (mis valkude järjestuse järgi aminohappe pos on määratud?) which are non-conserved between SARS-CoV and CoV-2 (**Supplementary Fig 2**). Ep.14 co-localizes with the fusion/entry domains of HR1 and Ep.15 is directly N-terminal to HR2 of S SARS-CoV-2^19^ (**Figure 1**).

**Figure 1.**
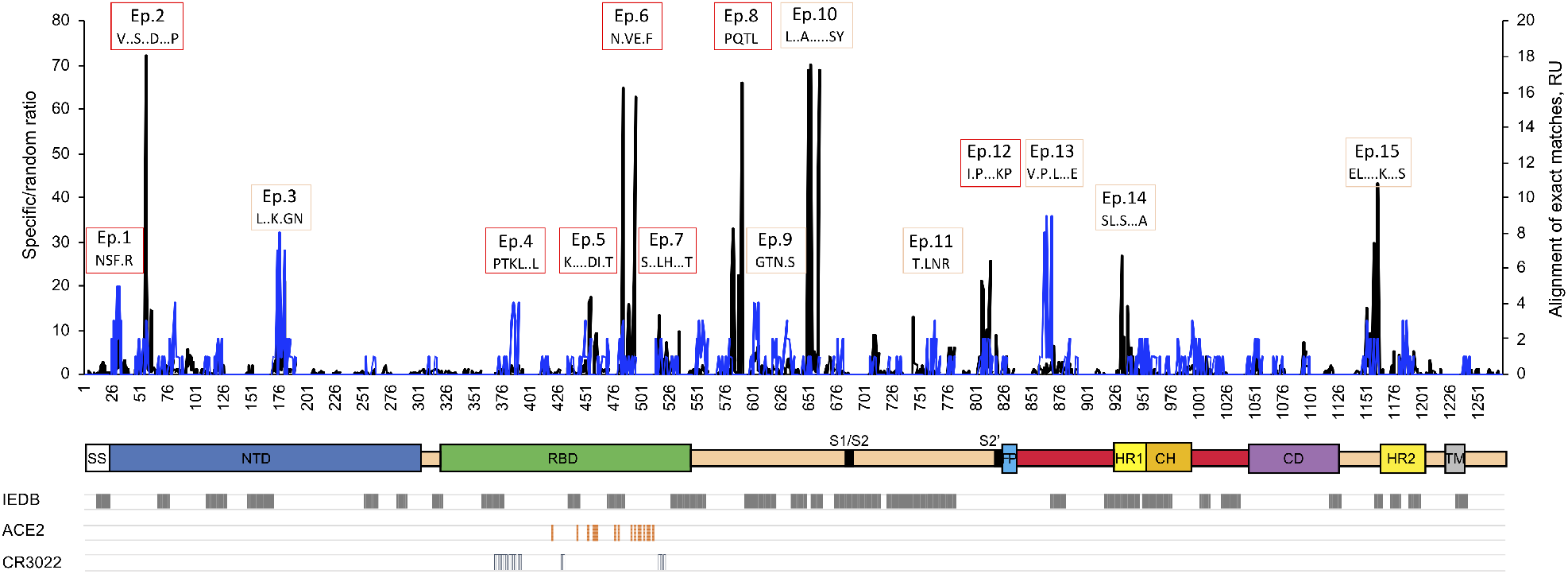
MVA predictions of epitopes for antibodies with known and novel specificity to S glycoprotein of SARS-CoV-2. Sequence alignment histograms for potential antibody-specificity against S reveals prominent epitopes with alignment loads above the 95% percentile. 22,949 unique core motif sequences (of 1,380,000 distinct peptide sequences), representing top most abundant and shared immune response, were screened for sequence similarity with SARS-CoV-2 Spike glycoprotein (P0DTC2) by alignment analysis. The specific/random ratio is the alignment of motifs matching with ≥4 amino acid positions (specific) in proportion to shuffled-sequence motifs (random). The ratio is calculated per each amino acid position on the primary sequence of S (*left y-axis, black line-graph*). Using simulated alignment distribution, 14 of predicted epitopes Ep.1 to Ep.15 were statistically significant (p-value < 0.05) (Ep.4 with p-value < 0.06). Of aligned motifs, 111 were found with exact-matching amino acid positions.The exact-matching motifs were counted for each amino acid position on primary sequence of S (*right y-axis, royal blue line-graph*)*. IEDB*, regions in *IEDB* that were predicted as B cell epitopes of S of SARS Cov-2^17^. MVA-predicted epitopes marked with *red empty boxes* differ from IEDB-marked epitopes. MVA-predicted epitopes marked with *rose gold empty boxes* overlap with IEDB-marked epitopes. *ACE2*, amino acid positions important for binding to human ACE2, the receptor for host cell infection^18^. *CR3022*, amino acid positions important for binding of CR3022 anti-SARS-CoV-2 Spike RBD antibody^18^. S glycoprotein domains are visualized schematically, adapted from Wrapp et al.^15^ with additional information about RBD from Yuan et al.^18^. SS – signal peptide (1-12); NTD – N-terminal domain (13-303); RBD – receptor binding domain (319-542); S1/S2 – S1 subunit end and S2 start site (683-686); S2’ – S2’ protease cleavage site (815-816); FP – fusion peptide (816-833); HR1 – heptad repeat 1 (908-985); CH – central helix (986-1035); CD – connector domain (1076-1141); HR2 – heptad repeat 2 (1163-1202); TM – transmembrane domain (1214-1234); RU – relative unit, count of motifs with exact identity to SARS-CoV-2 S in each amino acid position.

Next, we characterized the immunoreactivity patterns of the predicted epitopes of S across the whole cohort divided by different comorbidities and age (**Figure 2**). All examined epitopes, Ep.1 to Ep.15 exhibited immunoreactivity profiles that were highly individual-specific (**Figure 2A**). Exploratory data analysis revealed associations between immunoreactivity profiles and subject age, gender or clinical backgrounds (**Supplementary Table 2**), warranting more focused analysis.

**Figure 2.**
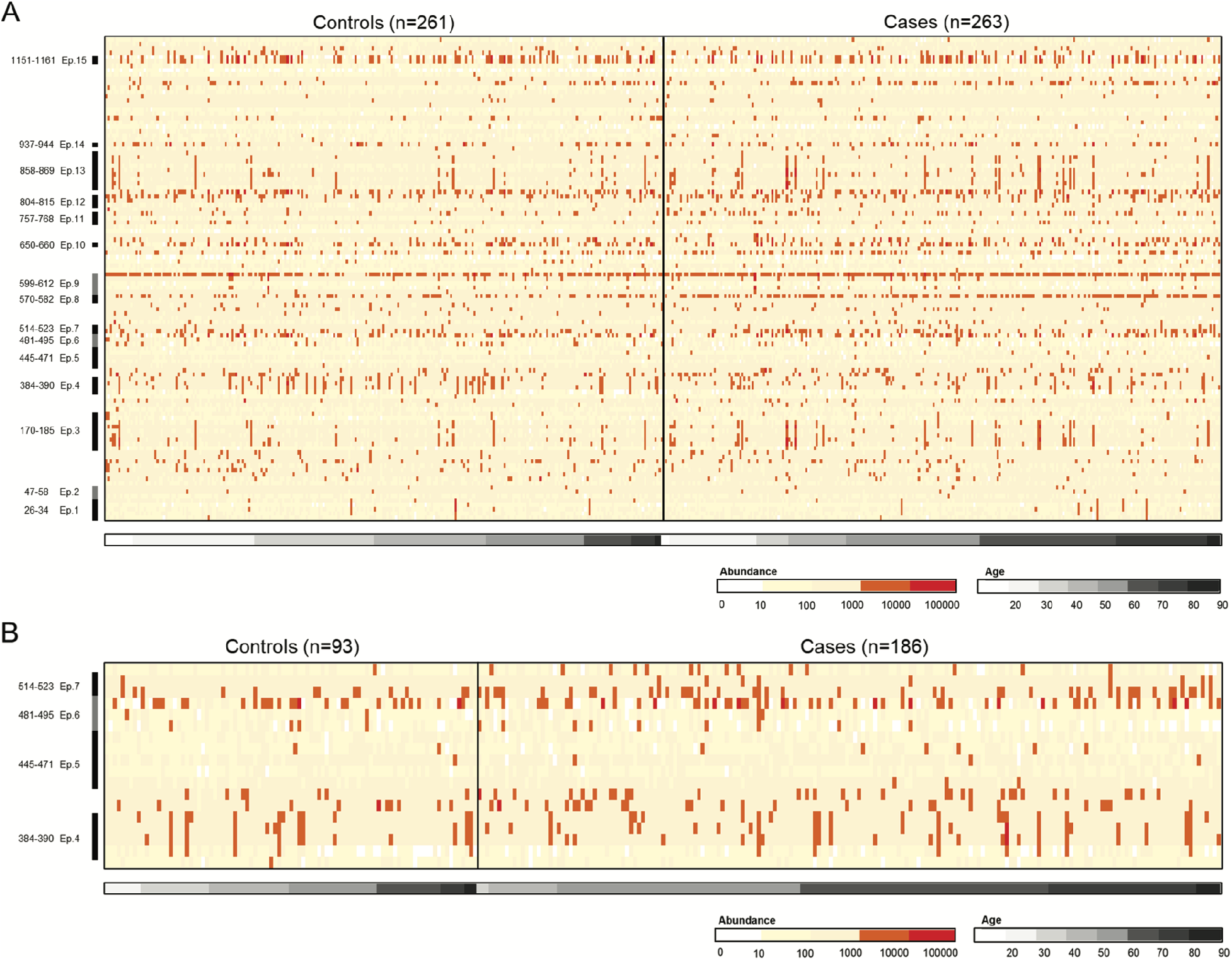
Immunoprofiles of peptide antigens with high similarity to the primary sequence of SARS-CoV-2 Spike glycoprotein (P0DTC2) across the study cohort (n=538) 22,949 unique core motif sequences (of 1,380,000 unique peptides), representing top most abundant and shared immune response, were screened for sequence similarity with SARS-CoV-2 spike glycoprotein P0DTC2 by alignment analysis. Of these, 111 were found across the primary sequence of Spike glycoprotein. **A |** The abundance of immunoprofile peptides containing the 111 motifs (*in rows*) was extracted for 538 samples, of which 524 were with known age (*in columns*) with varying clinical background, gender and age. The abundance of motif-containing peptides across subjects is visualized in log scale as intensity plot. The order of motifs is from N-terminal (down) to C-terminal (up) orientation on S, with predicted epitopes with spanning amino acids indicated on the left, where applicable. The result suggests high levels of individuality across predicted S-associated immunoprofile features. Peptide abundance (*abundance*) is color-coded in log_10_. Group abbreviations: Controls – healthy controls (HC groups), HC-MI, HC-CAD, HC-FEP, HC-SZ groups. Cases – MS – multiple sclerosis; BC – breast cancer; T2D – type 2 diabetes; MI – myocardial infarction; CAD – coronary artery disease; FEP – first episode psychosis; SZ – schizophrenia. **B |** Focused view at the clustering and abundance of epitope motifs on the receptor binding domain (RBD) of S (18 motifs in rows). Cases (n=186) included BC, T2D, MI, CAD, with matching controls where possible (n=93).

We did not detect any significant differences in neutralizing response (Ep.4 and Ep.7) among subjects older or younger than 61 years old (**Figure 2B** Kruskal-Wallis test, p>0.05 and **Supplementary Figure 3**). The abundance of antibodies predicted to interact with Ep.5 was higher among older subjects 61≤ than younger subjects aged 41-60 or ≤40 (Mann-Whitney U, ****p<0.0001) (**Figure 3A**). Intriguingly, Ep.5 is within the RBD of SARS-CoV-2 S protein^18^ (**Figure 1**) and contains overlapping key amino acids important for binding to ACE2 (**Supplementary Figure 2**). In Ep.5, the amino acids important for binding ACE2 are 100% conserved between SARS-CoV and CoV-2, with the exception of SARS-CoV-2 F456 -> SARS-CoV L443 transformation not targeted by aligned motifs (**Supplementary Figure 2**). Response against Ep.1 and Ep.8 of S was also found higher among older subjects aged 61 ≤ (Mann-Whitney U, **** p<0.0001) (**Figure 3A**). Response against Ep.6 of S was higher among younger subjects aged ≤40 or 41-60 than in subjects aged 61≤ (Mann-Whitney U, **** p<0.0001) (**Figure 3B**).

**Figure 3.**
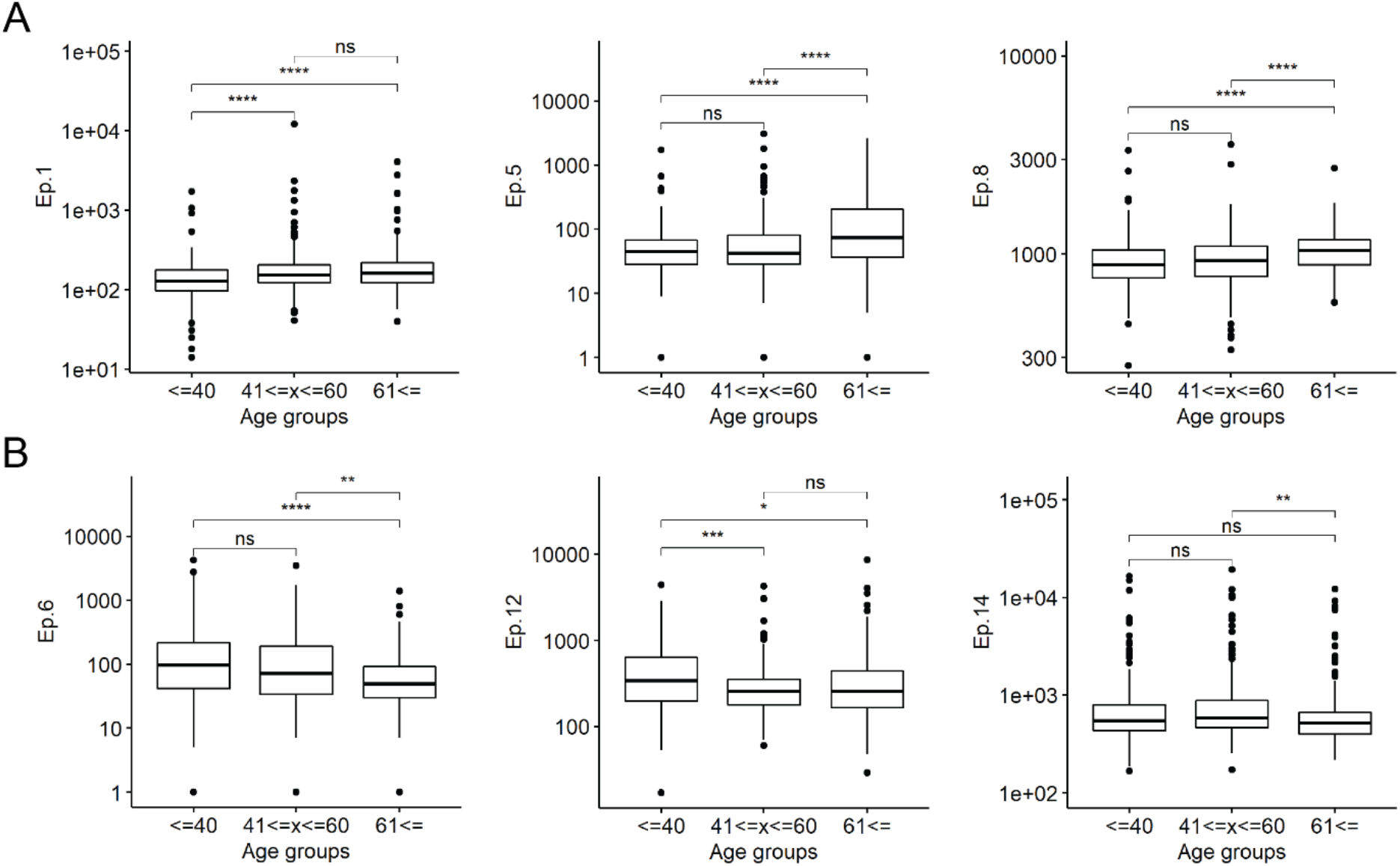
Age-associated differential antibody response to predicted epitopes of S RBD. The abundance of antibodies potentially specifically targeting S was assessed across age groups of 524 studied subjects. **A |** Antibody response to epitopes Ep.1, Ep.5 and Ep.8 of S was significantly higher in subjects aged 61≤ than in younger age groups. Pearson correlation for Ep.5 response (in log) and subject age was R = 0.20 (p=6.6 × 10-6, 95% CI (0.11, 0.28)), suggesting a weak positive correlation. Pearson correlation coefficient between subject age and Ep.1 response was R = 0.17 (p=1.3 × 104, 95% CI (0.08, 0.25)) and between anti-Ep.8 response R = 0.17 (p=6.2 × 10-5, 95% CI (0.09, 0.26)). Relative antibody response is assessed as abundance of peptide antigens containing epitope motif (*y-axis*) (**Figure 1**). Group sizes: ≤40 (n=186), 41≤x≤60 (n=188), 61≤ (n=150). **B |** Antibody response to epitopes Ep.6, Ep.12 and Ep.14 of S was significantly higher among younger ≤40 or 41-60 than in older subjects aged 61≤. Kruskal-Wallis test for multiple group comparison, pair-wise comparison with Mann-Whitney U test, ns p>0.05, * p<0.05, ** p<0.01, *** p<0.001, **** p<0.0001.

Antibody response against Ep.12 and Ep.14 of S2 of S was also statistically higher in younger subjects (Mann-Whitney U, * p<0.05, ** p<0.01, *** p<0.001) (**Figure 3B**), whereas response to remaining predicted S epitopes was not substantially different across age groups (**Supplementary Figure 3)**.

Next we analysed frequency of antibody responses to different epitopes in subjects with hypertension (HT), cardiovascular diseases (CVD) and subjects who smoked cigarettes. Ep.2, Ep.5, Ep.8, Ep.9, Ep.11, and Ep.13 were statistically (Mann-Whitney U) higher among subjects with HT (**Figure 4** and **Supplementary Figure 4** and **Figure 5**). An interesting difference was observed in male and female subjects with HT (**Figure 4** and **Supplementary Figure 6**). The higher response to Ep.5 of S RBD was preferentially in males with HT (Mann-Whitney U, * p<0.05, **Figure 4**), whereas the HT-associated higher response to Ep.8 was common to both sexes, however the male subjects with HT were found to have higher response to Ep.8 compared to female subjects with HT (**Figure 4**).

**Figure 4.**
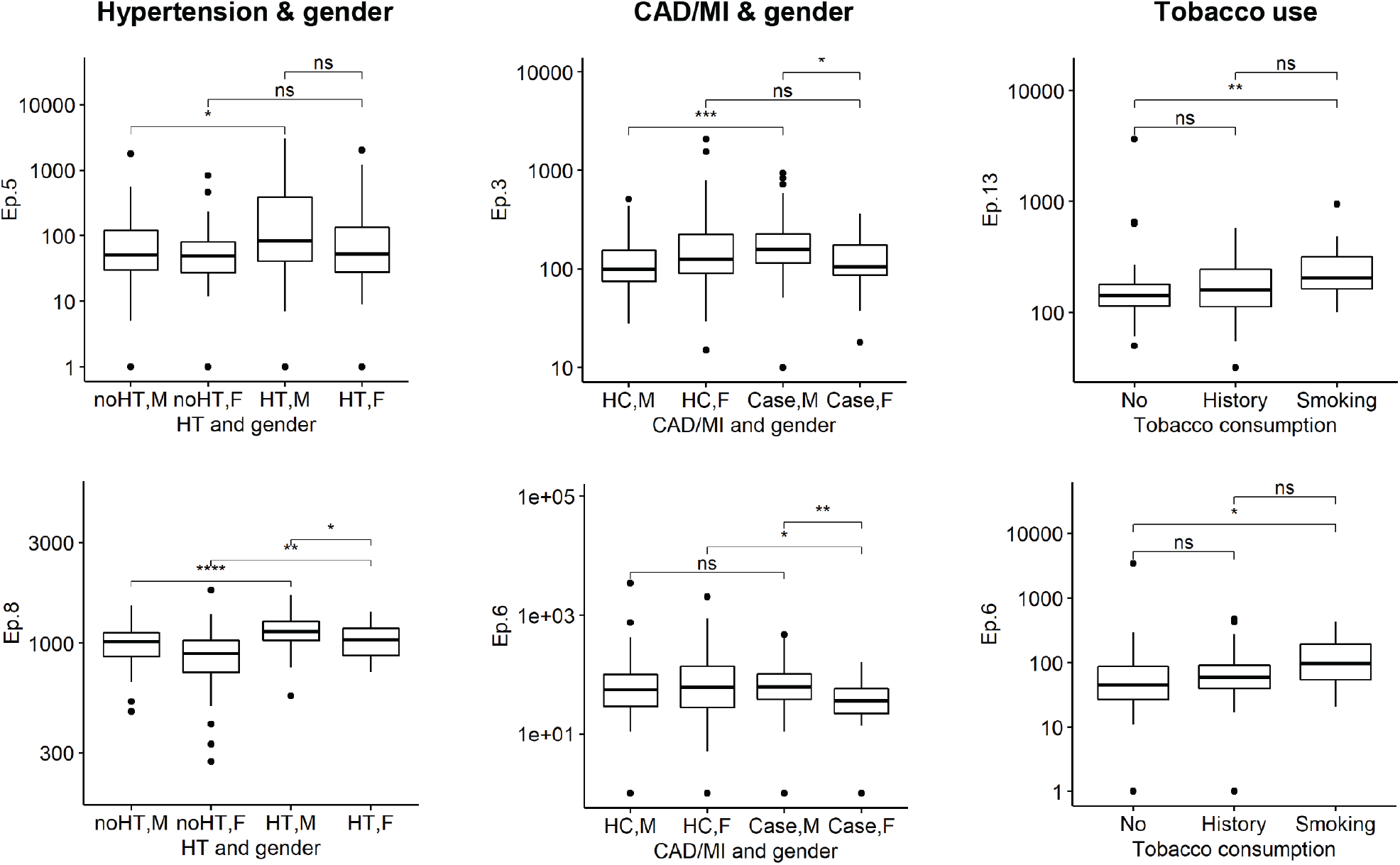
Specific higher response against epitopes of S was associated with hypertension (HT), cardiovascular diseases (CVD) and cigarette smokers. Statistical analysis identified anti-S response against epitopes of Ep.5 and Ep.8 as significantly higher in subjects with HT (n=112) than no-HT (n=81, **Supplementary Figure 5**). However, gender-based analysis revealed high Ep.5 response as male-HT specific and anti-Ep.8 response as higher in males with HT than in females with HT (**Supplementary Figure 6**). Relative antibody response is assessed as abundance of peptide antigens containing epitope motif (*y-axis*) (**Figure 1**). Group sizes: noHT, M (n=71), noHT, F (n=43), HT, M (n=48), HT, F (n=33). Male subjects with CAD or MI had higher response against Ep.3 of S than female subjects with CAD or MI (or healthy controls) (**Supplementary Figure 7)**. Female subjects with either CAD or MI showed reduced response against S Ep.6 than male subjects with CAD/MI (or healthy controls). Group sizes: HC, M (n=40), HC, F (n=49), Case, M (n=79), Case, F (n=27). Within the PD-CAD clinical cohort (n=96), subjects who were active cigarette smokers were identified as having higher response against predicted S Ep.6 or Ep.13 than non-smokers. Group sizes: Non-smokers (“No”) (n=49), previous history of smoking (“History”) (n=33), active cigarette smokers (“Smoking”) (n=14). Mann-Whitney U test, ns p>0.05, * p<0.05, ** p<0.01, *** p<0.001, **** p<0.0001. M, male, *F* female.

On the other hand, males with coronary artery disease (CAD) or MI showed statistically higher response to Ep.3 of S compared to females, whereas females with either CAD or MI showed significantly reduced response against Ep.6 of S compared to males (or healthy controls, **Figure 4, Supplementary Figure 7**). Cigarette smoking is a risk factor for both HT and CVD. However, some epidemiological observations suggest daily cigarette smokers have a lower probability of developing severe COVID-19 than the general population^20,21^. In the clinical cohort of CAD (n=96), predicted immune response to both Ep.6 and Ep.13 of S was identified as significantly higher among cigarette smokers than in non-smokers (**Figure 4**).

Taken together, our analysis predicited Ep.1 and Ep.3 (NTD of S), Ep.5 (RBD of S), Ep.8, and Ep.13 (S2 of S) as Spike epitopes linked to age, HT, CVD and tobacco use, all of which are confounding factors of COVID-19 severity. Although associated with tobacco use and gender-related cardiovascular condition, notably higher response to Ep.6 was correlated with younger age.

The observation of shared high immunogenicity of predicted epitopes of S suggested that regions in S contained evolutionarily and/or structurally related immunogenic targets. B cell-specific response potentially able to cross-react with S of SARS-CoV-2 might arise from exposure to other (beta-) coronaviruses commonly infecting humans^22^, or common human pathogen not evolutionarily related to coronaviruses but with protein structural similarity^14,23^. Sequence alignment analysis with common human beta-coronaviruses and across the human viral pathogenome (https://www.uniprot.org/help/uniprotkb) revealed substantial sequence similarity conservation of SARS-CoV-2 epitopes with other endemic CoVs (i.e. CoVs OC43 and HKU1) and other common pathogenic antigens, most notably human cytomegalovirus and human herpesvirus 6B (**Supplementary Figure 8, Table 3**), offering a mechanism for pathogenic cross-reactive mimicry. Exposure to many of the common human viruses usually occurs during childhood or adolescence that shapes our immune memory and modulates the subsequent response to self and environmental antigens later in life^9,10^.

The cell biology of SARS-CoV-2 is so far less known than that of other coronaviruses. Emerging evidence show that binding of SARS-CoV-2 to ACE2 on the surface of target cells such as lung endothelium is essential for viral infectivity. Our data reveal major epitope-specific responses to SARS-CoV-2 in an infection naïve population. Most interestingly, some of these antibodies overlap with crucial antigenic attachment region of SARS-CoV-2 S binding with ACE2, some with that of the neutralizing antibody (CR3022) of SARS-CoV-2, and some with cell fusion/entry domains HR1 and HR2. Our data also highlight the age-linked differential patterns of immune response to S, with individuals aged >60 being more apt to (suboptimal) antibody responses potentially affecting ACE2-S protein RBD of SARS-CoV-2 interactions (Ep.6, **Figure 1** and **3**).

These findings indicate that it is difficult to distinguish exposure to SARS-CoV-2 from other CoVs and common pathogens in serological studies using SARS-CoV-2 S as antigen. This implies that epitope-specific diagnostic assays will need to be designed. Our data further lends credence to the view that immunological memory covers families of antigens rather than a particular member only involved in the primary response, causing a rapid anamnestic response, at the expense of specificity^24,25^. These results also underscore the possibility of antigenic sin, shaping the immune response to SARS-CoV-2, which in turn could influence disease progression and the clinical outcome of COVID-19^14^.

## Methods

### Clinical cohorts

To investigate pre-existing immunoreactivity able to cross-react with the SARS-CoV-2 main antigenic epitopes on S protein, we examined the antibody immune profiles of several relevant clinical cohorts (**Table 1**). Blood samples were collected prior to SARS-CoV-2 emergence (2011 to September 2019) under local ethical approvals.

### Mimotope-variation analysis

For qualitative and quantitative characterization of humoral immune response from blood samples, we used an in-house developed mimotope-variation analysis (MVA) method as described previously^26^. In brief, a random 12-mer peptide phage library (Ph.D.-12, NEB, UK) was used according to the manufacturer’s protocol. Two μl of serum samples were incubated with 2.5μl library (~5 × 10^11^ phage particles) and immunoglubulin G (IgG) fraction was recovered by using protein G-coated magnetic beads (Thermo Fisher Scientific). Captured phage DNA was analysed by using next generation sequencing (Illumina HiSeq, 50-bp single end reads) with barcoded primers for sample multiplexing. To evaluate the reproducibility of proteomics data, we compared peptide abundance in two replicates using Pearson correlation coefficient, which was 0.985 (p<0.0001) (**Supplementary Figure 1**).

### Data analysis

Peptides in a sample dataset were trimmed to 3 million reads (RPM units). SPEXS2 exhaustive pattern search algorithm^26^ was used to cluster similar peptides and reveal recognition patters (epitope motifs) that would be enriched in studied peptide sets (Documentation: https://github.com/egonelbre/spexs2)). The identification of core motif sequences was either performed discriminatorily, where selected peptide sets were studied for enrichment compared to a control group, or non-discriminatorily, where enrichment was identified compared to a random-generated peptide set (hypothesis-free). In brief, for each clinical group, 100,000s of distinct peptide antigens were used as input, from which 1000s of distinct core epitope motifs were identified with SPEXS2 algorithm. The motifs were collected together to generate a broad screening set of 22,949 motifs, representing different age groups, genders, and clinical background. The 22,949 motif sequences were aligned to primary sequence of SARS-CoV-2 Spike glycoprotein (S) (UniProtKB accession P0DTC2). A detailed description for clustering workflow is in **Supplementary Information**.

### Sequence alignment

SARS-CoV-2 proteome and Spike (S) glycoprotein (P0DTC2) were obtained from www.viralzone.expasy.org/8996 (date accessed 25.03.2020). To predict immunogenic regions on S, in-house built custom algorithms were used for sequence alignment. Primary sequence of S was scanned with 22,949 distinct motifs with criterium of ≥4 exact-matching amino acid positions. Random reference profile was generated by scanning with shuffled-sequence motifs (≥4 exact positions). Random alignment distribution was simulated with 3 independent alignments with shuffled motifs. Specific alignments above 0.95 percentile of simulated distribution were statistically significant (p<0.05). Alignment profiles were visualized using custom Excel VBA scripts and MS Office Excel.

Predicted B cell epitopes for the SARS-CoV-2 virus were published by Grifoni and colleagues^17^. Amino acids on Spike glycoprotein (S) of SARS-CoV-2 important for binding to human ACE2 were recently reported by different groups and anti-Spike RBD neutralizing antibody CR3022 discontinuous epitope were reported by Yuan and colleagues^18^. Domains of S were accessed from recently published data^15,18^.

SARS-CoV S glycoprotein primary sequence (P59594) was accessed from UniProtKB (date accessed 25.03.2020). To assess conservation of S from SARS-CoV-2, SARS-CoV, OC43 and HKU1 sequence alignment was performed with Clustal Omega tool (version 1.2.4) with default settings (https://www.ebi.ac.uk/Tools/msa/clustalo/).

Reference proteins for SARS-CoV-2 Spike glycoprotein (S) alignment with common human pathogens were obtained from UniProtKB with keywords “human” + “virus” (date accessed: 16.12.2019). The primary sequences of reference proteins, were first scanned with 111 SARS-CoV-2 S protein motifs with the criterium of an exact match. The reference proteins, which gave a match with S protein motif alignment, were consequently scanned with predicted S protein epitope sequences with the criteria of ≥5 exact-matching amino acid positions. Alignment procedures were carried out using custom Excel VBA scripts and MS Office Excel. Known epitopes of reference proteins that showed potential for cross-reactions with SARS-CoV-2 S protein were obtained from Immune Epitope Database’s full epitope list (epitope_full_v3.zip) (accessed 29.04.2020).

### Statistical analysis

All statistical analyses (ANOVA, t-Test, correlation analyses, ROC analysis) were done using R statistical programming language and RStudio environment (RStudio Team, 2015; R Core Team, 2017). Boxplot graphs were produced and visualized using packages “reshape2”, “dplyr”, and “ggplot2” 2020 versions ^27^. For visualization of peptide abundance across the sample set, peptide frequencies were converted to base 10 logarithmic values and intensity plots were graphed in MS Office Excel.

## Data Availability

The data that support the findings of this study are available on request from the corresponding author (KP). The data are not publicly available due to containing information that could compromise research participant privacy/consent. Any materials that can be shared will be released via a material transfer agreement.

## Ethics Statement

Immunoprofiling of the samples has been approved by the institutional ethics committees, and all subjects provided written informed consent.

## Author Contributions

AR and NP contributed equally to the work as 2nd authors. KP, MJ, AR, NP, and AMG conceived and designed the study. AP, MP, LH, EV, MB, FL, EK, PP, PT, AV and DL collected samples and acquired the clinical data. KP, MJ, AR, NP, AP, HS, AA and AMG analyzed and interpreted the data. EV, PT, DL, TT and KP supervised the study. KP and AMG drafted the manuscript. All authors revised and approved the final manuscript for submission.

## Acknowledgments

Mirjam Luhakooder, Helen Verev, Susan Pihelgas and Maila Rähn are thanked for excellent technical assistance.

